# Aggregating Multiple Real-World Data Sources using a Patient-Centered Health Data Sharing Platform: an 8-week Cohort Study among Patients Undergoing Bariatric Surgery or Catheter Ablation of Atrial Fibrillation

**DOI:** 10.1101/19010348

**Authors:** Sanket S. Dhruva, Joseph S. Ross, Joseph G. Akar, Brittany Caldwell, Karla Childers, Wing Chow, Laura Ciaccio, Paul Coplan, Jun Dong, Hayley J. Dykhoff, Stephen Johnston, Todd Kellogg, Cynthia Long, Peter A. Noseworthy, Kurt Roberts, Anindita Saha, Andrew Yoo, Nilay D. Shah

## Abstract

Real-world data sources, including electronic health records (EHR) and personal digital device data, are increasingly available, but are often siloed and cannot be easily integrated for clinical, research, or regulatory purposes. We conducted a prospective cohort study of 60 patients undergoing bariatric surgery or catheter-based atrial fibrillation ablation at two U.S. tertiary care hospitals, testing the feasibility of using a patient-centered health data sharing platform to obtain and aggregate health data from multiple sources. We successfully obtained EHR data for all patients at both hospitals, as well as from 10 additional health systems, which were successfully aggregated with pharmacy data obtained for patients using CVS or Walgreens pharmacies, personal digital device data from activity monitors, digital weight scales, and single-lead ECGs, and patient-reported outcome measure data obtained through surveys to assess post-procedure recovery and disease-specific symptoms. A patient-centered health data sharing platform successfully aggregated data from multiple sources.

## INTRODUCTION

Medical products, including drugs and devices, play an important role in clinical medicine, can provide substantial benefits to patients, and are regulated by the Food and Drug Administration (FDA) in the United States. In recent years, FDA increasingly moved towards a total product life cycle approach to medical product oversight, particularly for medical devices, with increasing use of evidence derived from real-world data collected in the post-market setting as part of efforts to further evaluate medical product safety and effectiveness.^1,2^ Real-world data used for this purpose can be derived from multiple sources, such as administrative claims, electronic health records (EHRs), disease and device registries, data gathered through personal digital devices, and patient-generated health data;^2-4^ however, aggregating data from multiple sources is often challenging.

Real-world data sources are most often siloed and represent only specific aspects of a patient’s experience. For example, one health system might have data from its EHR and some patient-generated health data, but because of lack of interoperability, is unlikely to have access to data from EHRs of other health systems where a given patient receives care. Similarly, registries extensively curate health data for patients, generally organized around patients having been diagnosed with a specific disease or receiving a specific device or procedure. But these data are typically limited to a point in time, focused on disease or device-specific data elements, and often have limited follow-up. When the Health Information Technology for Clinical and Economic Health (HITECH) Act was enacted in 2009, a key provision was for healthcare providers to provide people with digital access to their health records. Known as the Blue Button project, this effort enabled patients to access their personal health information as digital data through patient portals,^5^ facilitating individual agency over their data, and has since been extended to administrative claims.^6^ While portals provide patients access to their data, separate portals are often required if they receive care in different healthcare systems.

Mobile health (mHealth) technologies present an opportunity to obtain and aggregate multiple types of health data from multiple sources (e.g. EHR data from multiple healthcare systems), leveraging Blue Button technology or Application Programming Interfaces (APIs). Recent studies have shown that patients can be quickly enrolled into mHealth applications that allow them to contribute their personal digital data to research studies.^7^ Additionally, mHealth technologies can enable people to answer questionnaires about their health and contribute patient-reported outcome measure (PROM) data; PROMs have been shown to correlate with digitally-obtained physical activity levels^8^ and clinician-assessed performance status.^9^ However, to our knowledge, no study to date has aggregated multiple sources of patient health data, including personal digital device data, PROM data, EHR data, and data from pharmacies, devising an integrated approach that provides patients agency over their data while enabling pragmatic clinical research. Obtaining and rapidly aggregating these different real-world data sources for patients receiving therapeutic medical devices or procedures could help advance our understanding of medical device safety and effectiveness and provide a patient-centered approach to generating real-world evidence.

Accordingly, we conducted a study to test the feasibility of using a novel patient-centered health data sharing platform, Hugo (Hugo Health; Guilford, CT), to aggregate multiple real-world data sources as part of a prospective cohort study. Hugo facilitates patients’ access to their EHR, pharmacy, and patient-generated health data from personal digital devices (such as wearable and sensor-enabled devices) on any personal mobile device. Importantly, Hugo can aggregate data from multiple healthcare systems, wherever a patient receives care. In addition, if patients use and sync their personal digital devices, these data, along with those from EHRs and pharmacies, can be obtained at pre-specified intervals that can be near real-time. Patients can also elect to receive survey questionnaires to assess PROMs and other measures of patient status and symptom burden, responses from which are made available as soon as they are completed. Hugo allows patients to share these data with researchers through a permission-based system for research purposes. We conducted an 8-week cohort study to assess the ability of the Hugo mHealth platform to aggregate multiple sources of real-world and healthcare system data for patients undergoing two procedures that use medical devices: bariatric surgery (specifically, sleeve gastrectomy and gastric bypass) and catheter-based atrial fibrillation ablation.

## METHODS

### Study Population and Enrollment

We enrolled a convenience sample of patients who were planning on undergoing either bariatric surgery (sleeve gastrectomy and gastric bypass) or catheter-based atrial fibrillation ablation at Yale-New Haven Hospital (YNHH) or the Mayo Clinic. All patients had to be older than 18 years, English-speaking, have a compatible smartphone or tablet, and either have an email address or agree to create one.

Eligible patients were identified by their treating bariatric surgeon or cardiac electrophysiologist in advance of or following their final pre-procedural appointment. These patients were then contacted by phone and informed about the opportunity to participate in a digital health study to test the feasibility of using a novel mHealth application to enable integration of their data post-procedure. They were informed that this study would neither impact their peri-procedural care nor standard follow-up, as all data were collected only for research purposes to test the feasibility of the platform and were not related to the direction of their treatment. Study coordinators at both sites then met with patients to describe the study in full, obtain consent (from those who were willing), and enroll them into the mHealth platform, Hugo. Patients were offered a stipend to cover time and effort involved with study enrollment and participation.

Enrollment began on January 26, 2018 and continued until 15 patients were enrolled for each procedure at each site, for a total of 60 patients. Patients were followed for 8 weeks after their procedure. This study received Institutional Review Board approval at Yale University and Mayo Clinic. An IRB Authorization Agreement was signed by FDA. This study was registered at Clinicaltrials.gov (NCT03436082).

### Data Sources Aggregated by the Hugo Platform

We tested the feasibility of the Hugo platform to aggregate 4 different sources of data: electronic health records, data from pharmacies, patient-generated health data from personal digital devices, and PROM data (Figure 9).

**Figure 1:**
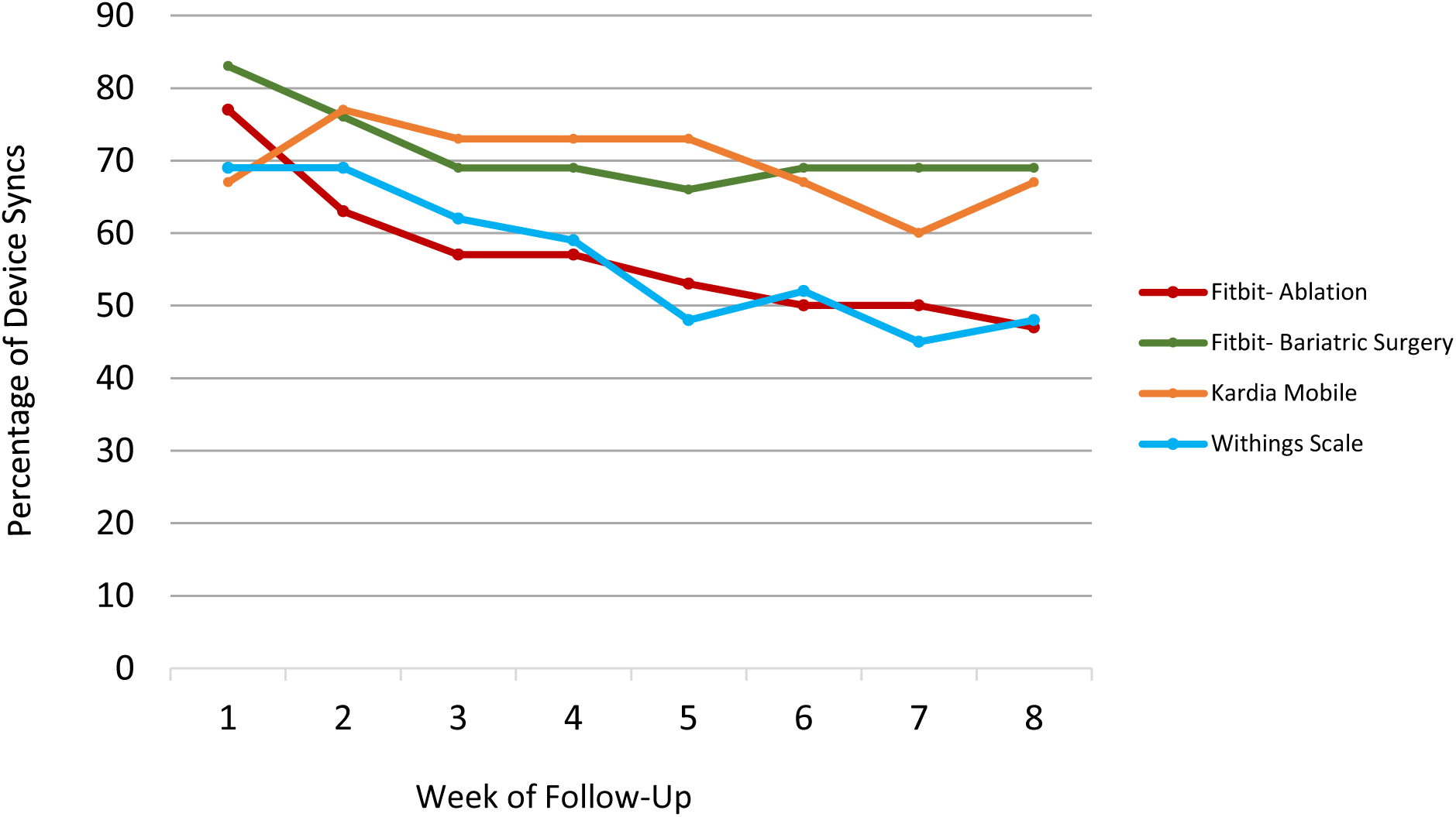
Rates of Weekly Personal Digital Device Use and Syncing over 8-Week Follow-Up Period among Patients after Bariatric Surgery or Atrial Fibrillation Ablation (n=59)

**Figure 2:**
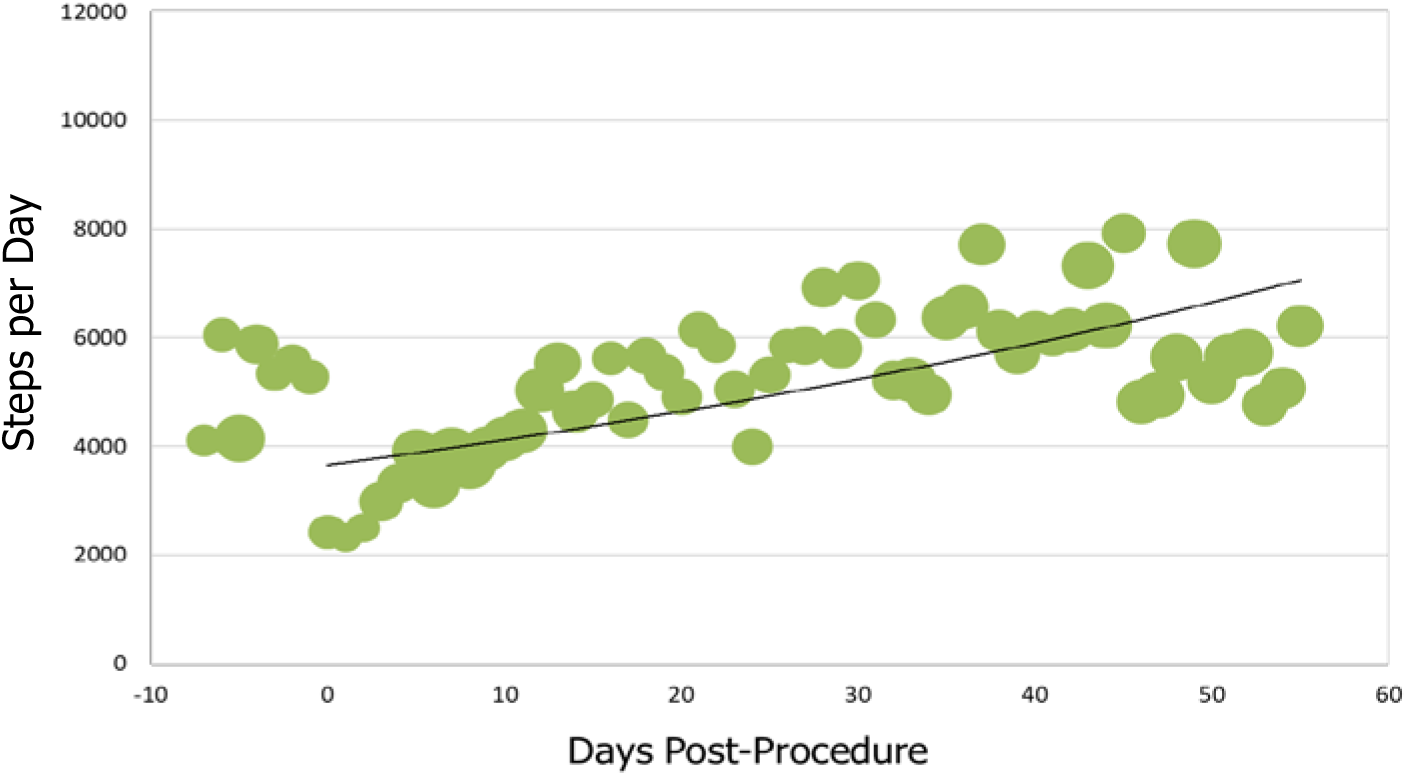
Average Steps Per Day for Patients who Underwent Bariatric Surgery (n=29). Bubble size corresponds to number of patients for whom data were synced. Procedure occurred on Day 0. Negative days indicates pre-procedure data, when available. Data obtained via Fitbit Charge 2

**Figure 3:**
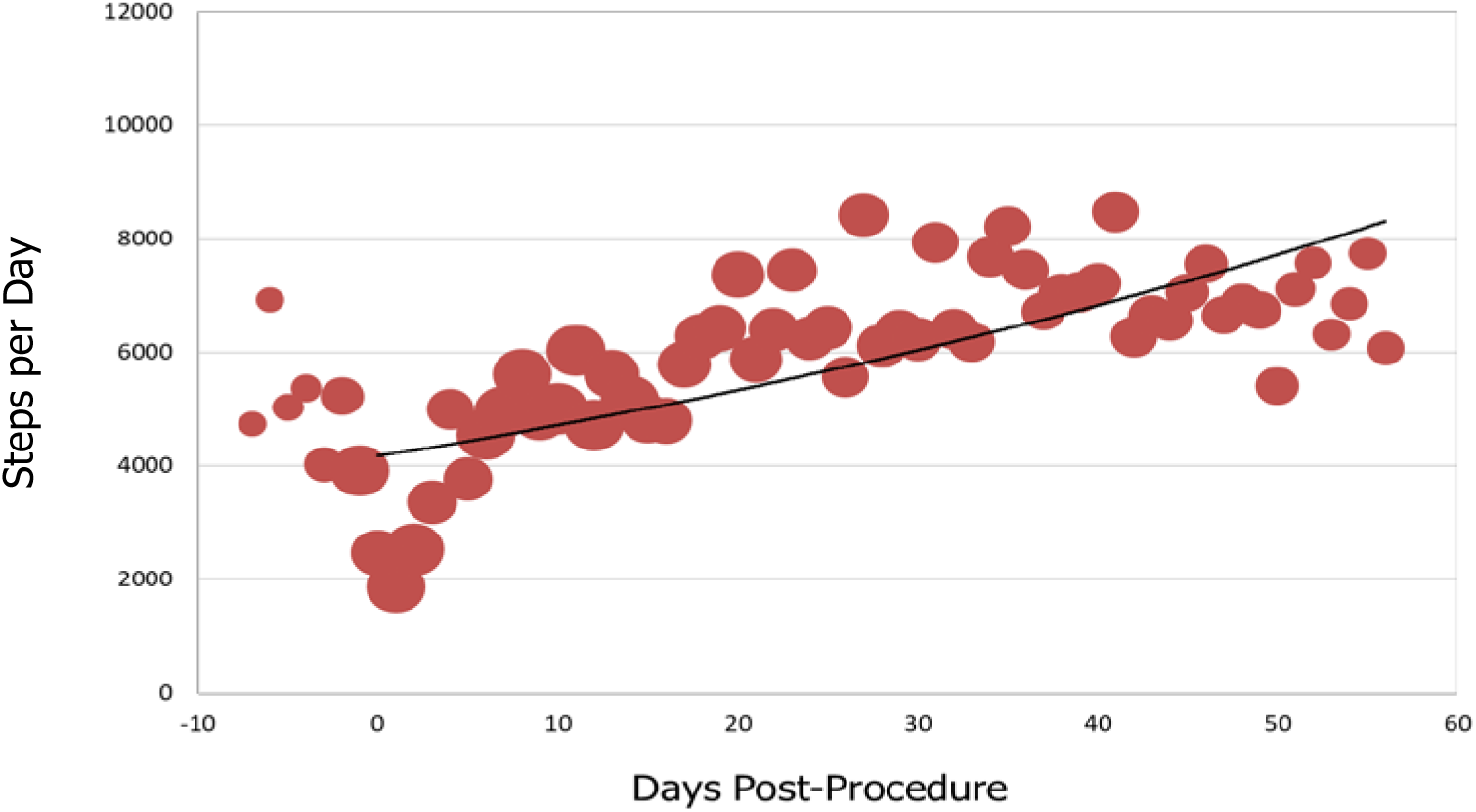
Average Steps Per Day for Patients who Underwent Atrial Fibrillation Ablation (n=30). Bubble size corresponds to number of patients for whom data were synced. Procedure occurred on Day 0. Negative days indicates pre-procedure data, when available. Data obtained via Fitbit Charge 2

**Figure 4:**
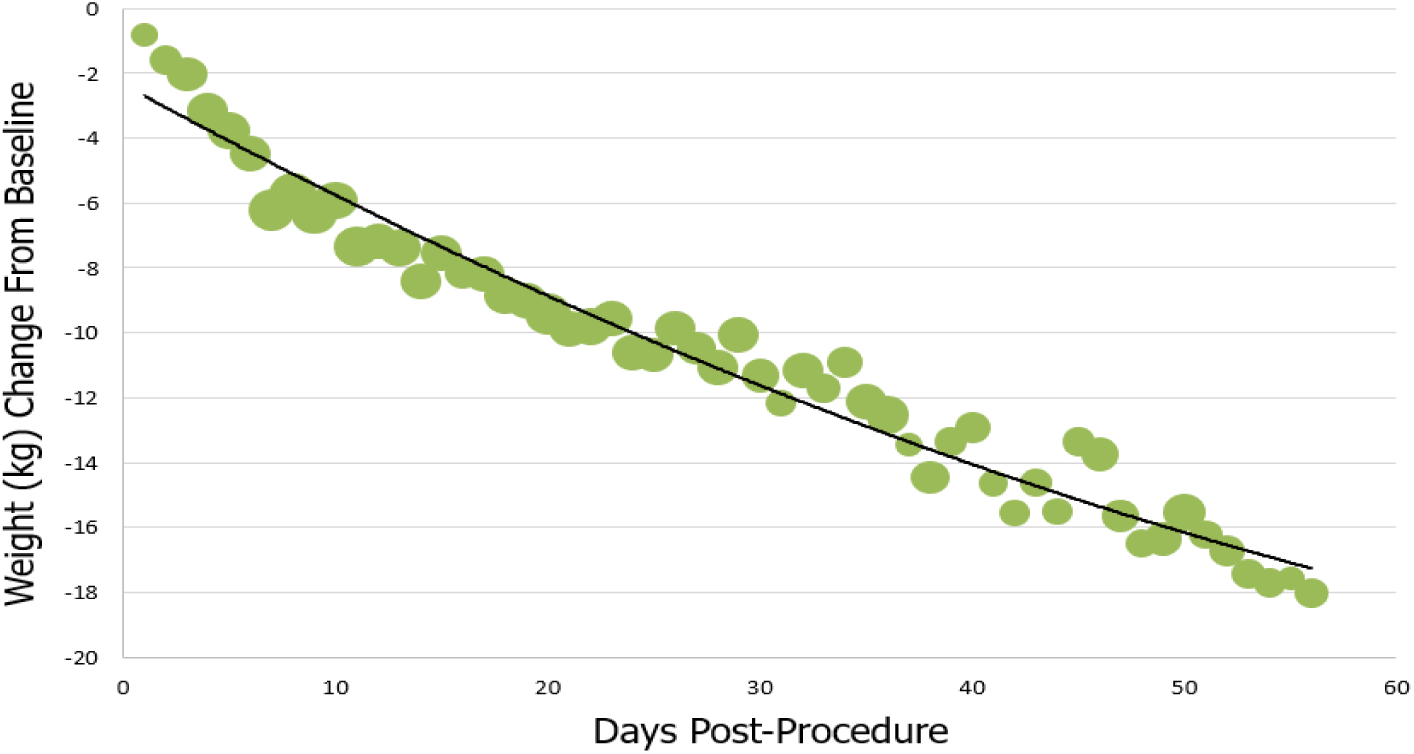
Average Cumulative Weight Change from Baseline for Patients who Underwent Bariatric Surgery (n=29). Bubble size corresponds to number of patients for whom data were synced. Procedure occurred on Day 0. Data obtained via Withings Body Scale

**Figure 5:**
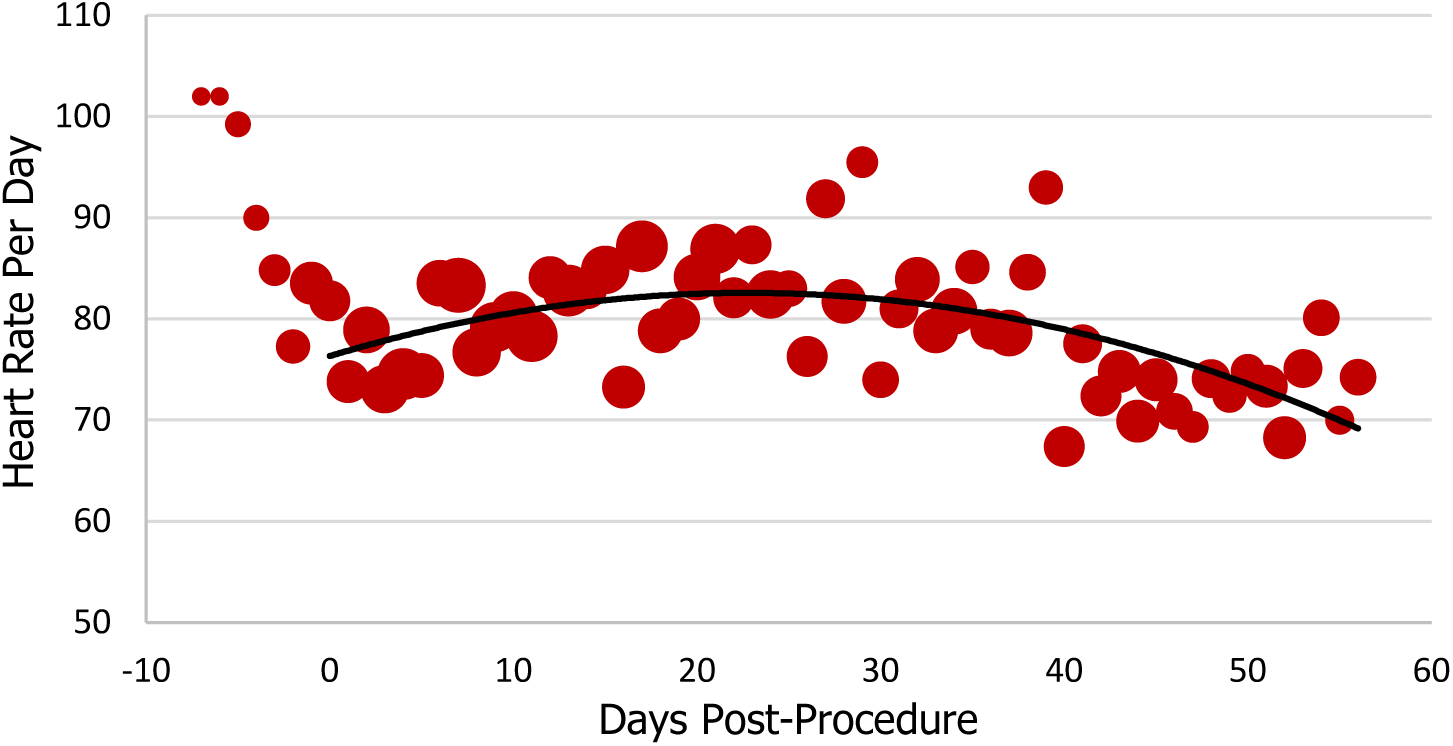
Average Heart Rate Per Day for Patients who Underwent Atrial Fibrillation Ablation (n=30). Bubble size corresponds to number of patients for whom data were synced. Procedure occurred on Day 0. Negative days indicates pre-procedure data, when available. Data obtained via AliveCor Kardia Mobile

**Figure 6:**
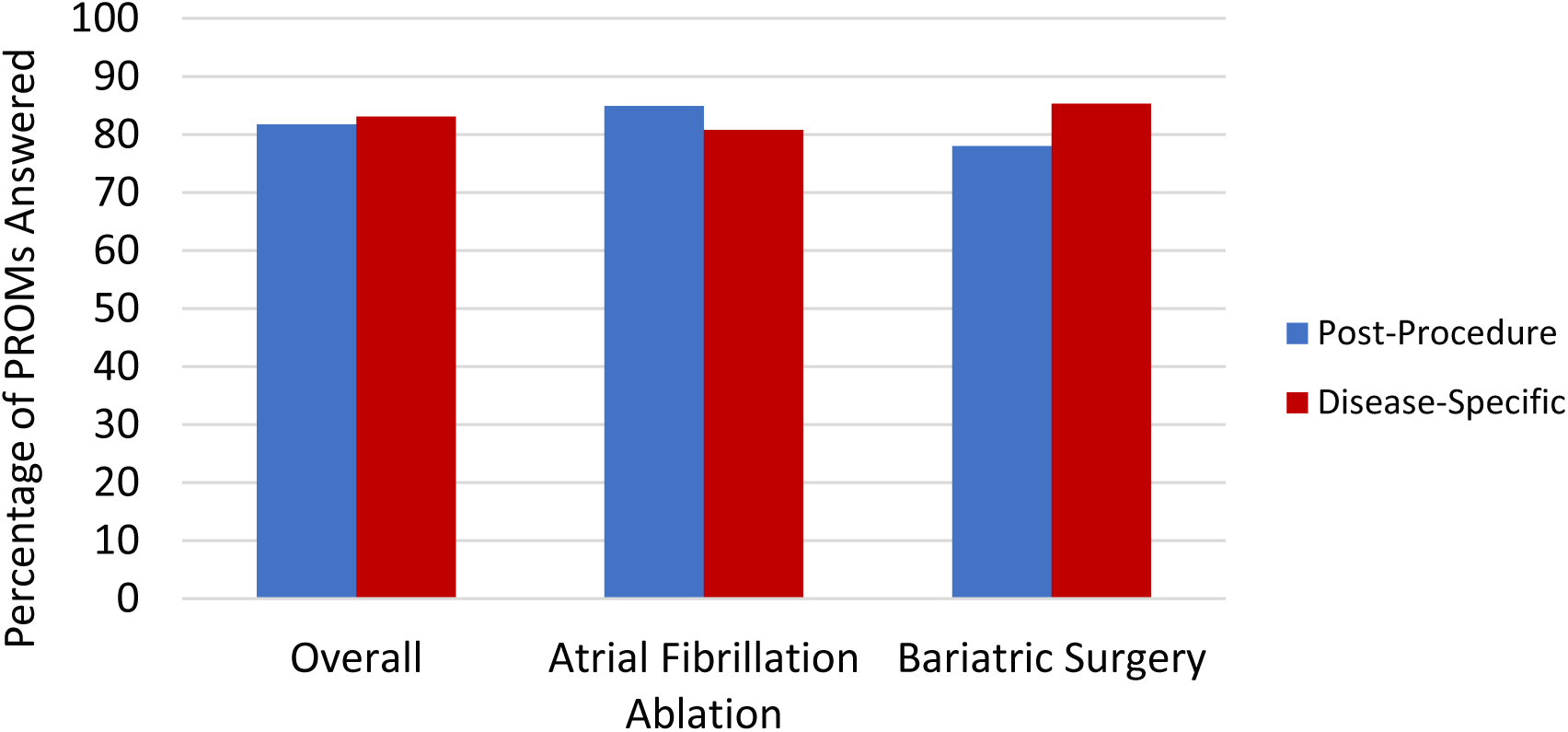
Percentage of Post-Procedure Recovery and Disease-Specific Patient-Reported Outcome Measure Questionnaires Answered.

**Figure 7:**
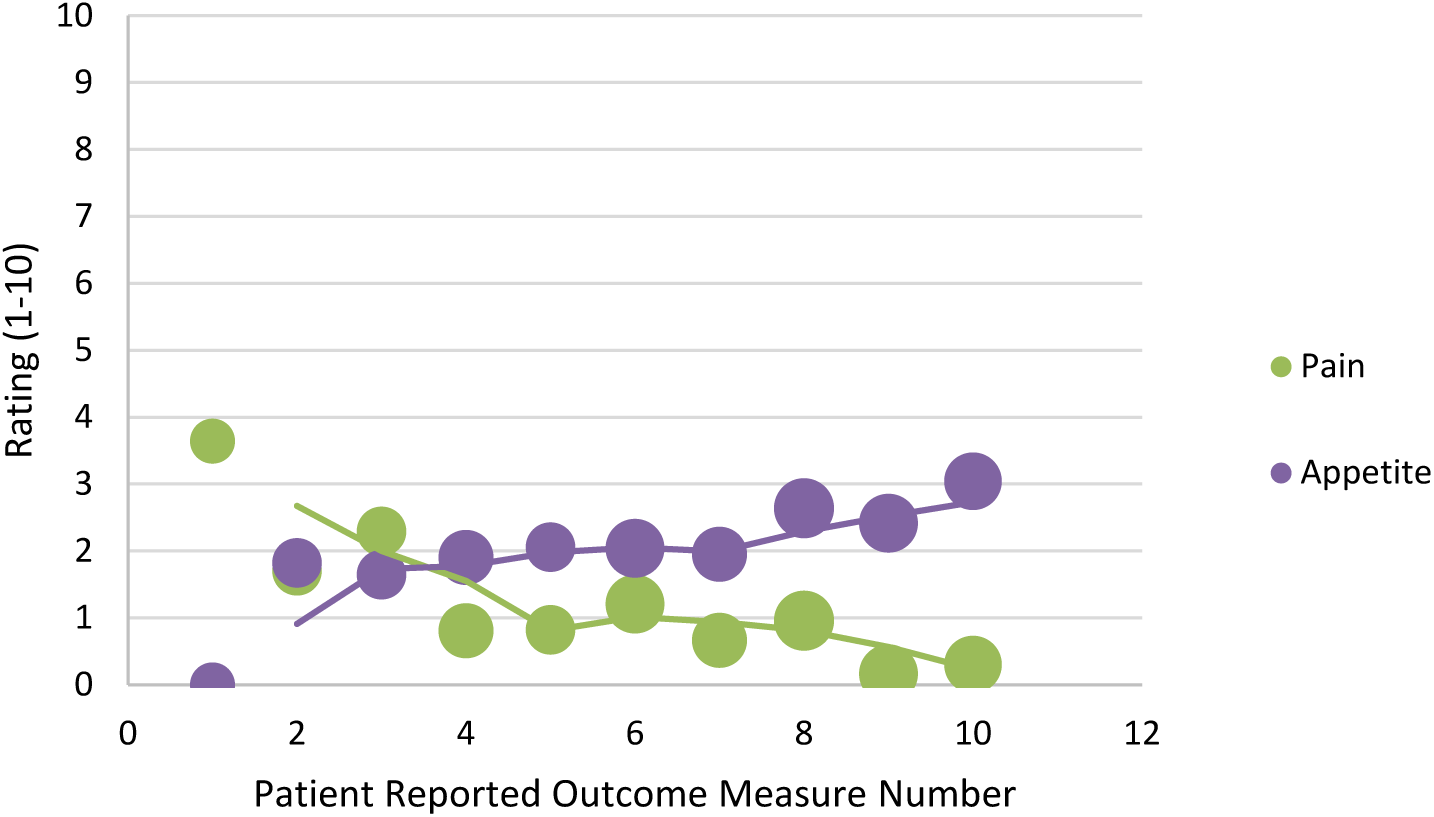
Responses to Post-Procedure Recovery Patient-Reported Outcome Measure Questionnaires about Pain and Appetite Among Post-Operative Bariatric Surgery Patients. Post-Procedure Patient Reported Outcome Measure Questionnaires were sent twice weekly, on Mondays and Thursdays, for a total of 10 instances post-procedure. Regarding pain, patients were asked, “Do you have any pain (yes/no)?” and if they answered yes, were asked “How would you rate your pain?” (1 being mild, 10 being severe on a visual analog scale). Regarding appetite, patients were asked, “Do you have an appetite (yes/no)?” and if they answered yes, were asked “How strong is your appetite?” (1 being weak, 10 being strong on a visual analog scale). Bubble size corresponds to number of patients for whom patient reported outcome measure questionnaire data were available

**Figure 8:**
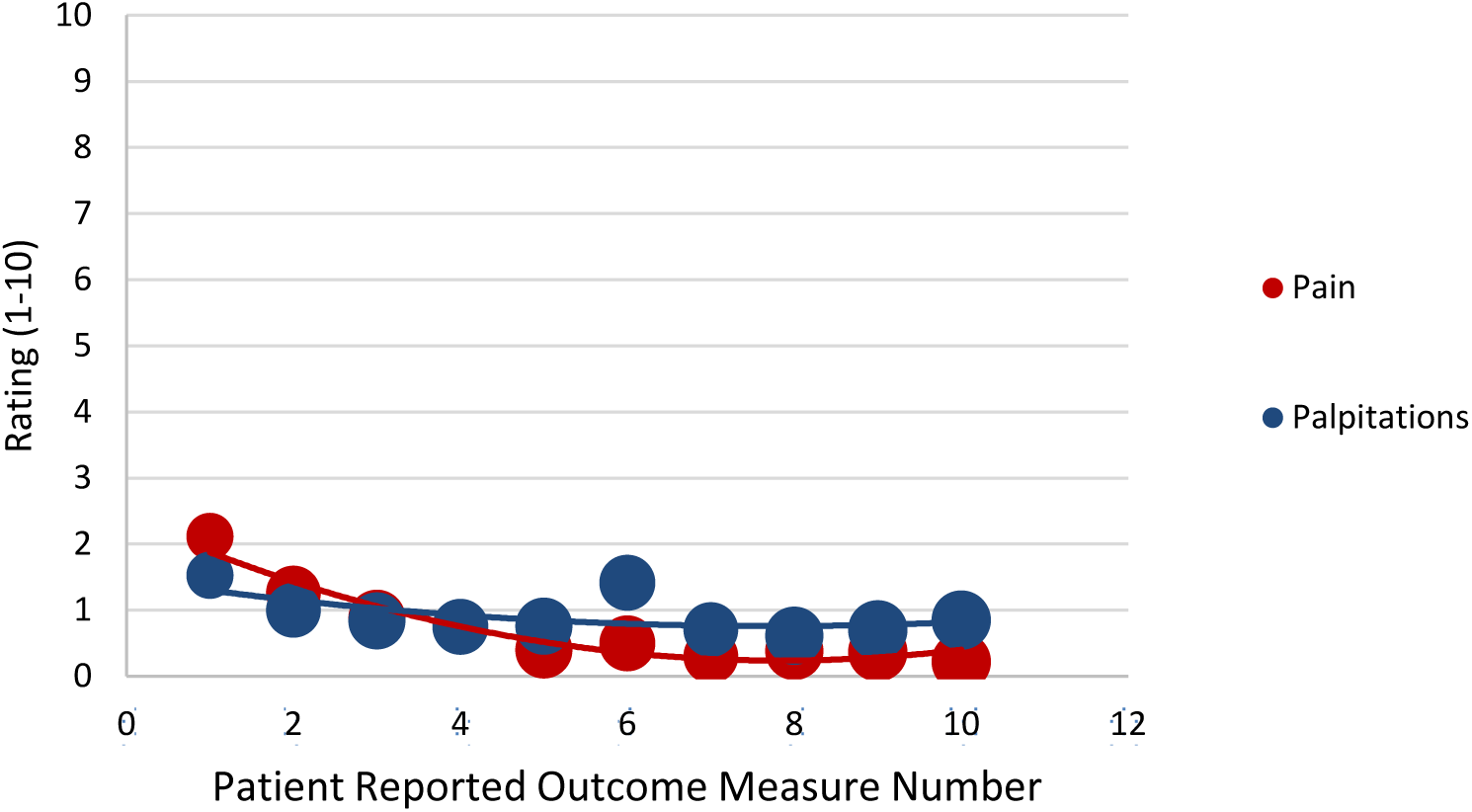
Responses to Post-Procedure Recovery Patient-Reported Outcome Measure Questionnaires about Pain and Palpitations among Patients Undergoing Atrial Fibrillation Ablation. Post-Procedure Patient Reported Outcome Measure Questionnaires were sent twice weekly, on Mondays and Thursdays, for a total of 10 instances post-procedure. Regarding pain, patients were asked, “Do you have any pain (yes/no)?” and if they answered yes, were asked “How would you rate your pain?” (1 being mild, 10 being severe on a visual analog scale). Regarding palpitations, patients were asked, “Do you have any palpitations (yes/no)” and if they answered yes, were asked “How would you rate your palpitations?” (1 being mild, 10 being severe, on a visual analog scale). Bubble size corresponds to number of patients for whom patient reported outcome measure questionnaire data were available

**Figure 9:**
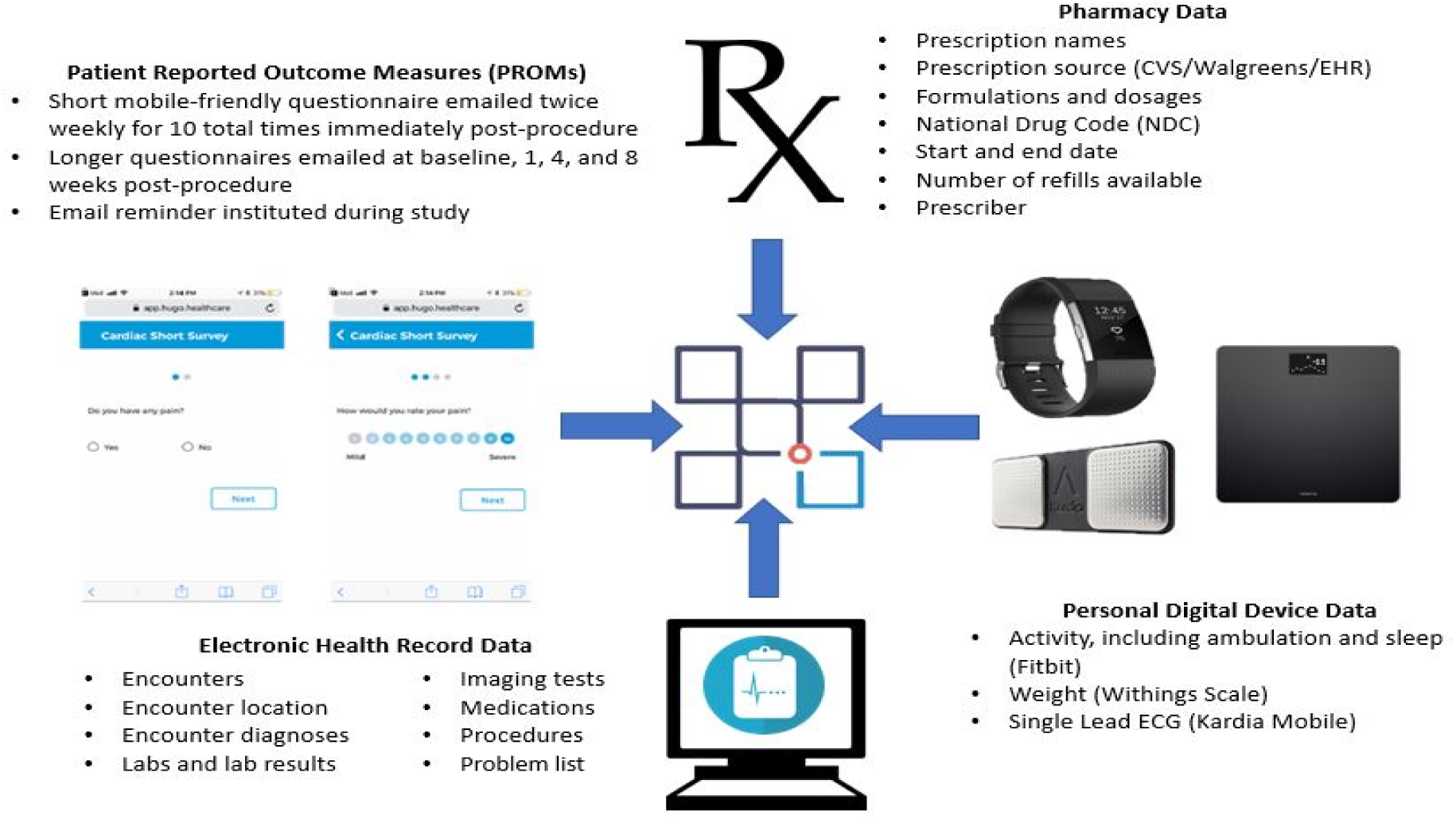
Data Aggregated in the Hugo Platform. Devices pictured include Fitbit Charge 2, Withings Body Scale, and AliveCor Kardia Mobile

#### Electronic Health Records

Hugo aggregates EHR data by having patients link their health system patient portals. At the time of enrollment, Hugo was connected to more than 500 health systems using either an Epic or Cerner EHR system. Hugo can integrate with additional United States EHR vendors. Through this linkage, patients obtain access to their CCD from each of the health systems where they receive care. The content of CCDs varies by health system, but generally includes encounter dates, encounter types, encounter diagnoses, medications, problem list items, lab results, and imaging test results. Hugo enables patients to share the data within the CCD with researchers. The Hugo team then extracted data from the CCDs to provide a more user-friendly .csv format for our research team.

All study patients recruited from YNHH were asked to link their patient portals to the Hugo application by entering their patient portal credentials (username and password). When necessary, our study coordinators assisted patients with creating patient portal accounts. Additionally, patients were asked to link portals from any other health systems where they received care (i.e., outside of YNHH and Mayo Clinic). If a specific health system portal was unavailable within Hugo, this was documented by the study coordinator and shared with the Hugo team to determine whether that health system could be connected to the Hugo platform. Because Mayo Clinic was amidst a transition in EHR vendors during the time of the study, we did not use the standard Hugo mechanism of having patients connect their Mayo Clinic portals. Instead, for patients recruited from Mayo Clinic, every week each patient’s updated CCD as proxy EHR data was sent directly to Hugo, where they were uploaded and made available to patients through the Hugo application.

#### Pharmacy

At the time our study was conducted, the only large pharmacy chains in Connecticut and Minnesota making their data available through patient portals were CVS and Walgreens. Walmart’s technical systems were in transition. Hugo aggregates pharmacy data from CVS and Walgreens by having patients link their pharmacy portal in a similar fashion as their patient portal. All patients recruited from either YNHH or Mayo Clinic who used either CVS or Walgreens as their primary pharmacy were asked to link their portals. When necessary, our study coordinators assisted patients in creating new pharmacy portal accounts. This linkage enabled Hugo to aggregate patients’ prescription medication names, dosages, instructions, national drug code (NDC), prescriber name, start date, and number of refills remaining.

#### Patient-Generated Health Data: Personal Digital Devices

Hugo connects to the public API for various personal digital devices. To collect personal digital device data over the course of the study, all patients received a Fitbit Charge 2 (Fitbit, San Francisco, California). Patients undergoing bariatric surgery also received a Withings digital weight scale (Withings, France), and patients undergoing atrial fibrillation ablation received a Kardia Mobile device (AliveCor, Mountain View, California). Study coordinators guided patients through device set-up at enrollment, including linking the devices to patients’ Hugo accounts, allowing Hugo to aggregate the information from each of the personal digital devices. After enrollment, patients received a follow-up email with user guides for the devices. Patients were asked to sync these personal digital devices at least once weekly during the 8-week post-procedure period. Beginning on July 6, 2018, approximately two-thirds through study completion, Hugo initiated a single weekly email reminder to remind all patients to utilize and sync their connected devices.

The data elements captured by a Fitbit Charge 2 included daily step counts and sleep duration. In some cases, physical activity, including the activity type and calories burned, was obtained automatically or entered by patients. Patients could also manually enter their height and weight. The sole data element captured by the Withings scale was weight, although patients were able to manually enter their heart rate. The data elements captured by the Kardia Mobile included reading duration, heart rate (usually averaged over 30 seconds), and a rhythm interpretation (atrial fibrillation, normal, or undetermined).

#### Patient-Reported Outcome Measures (PROMs)

Hugo allows patients to elect to receive survey questionnaires to assess PROMs and other measures of patient status and symptom burden through emails that generate a secure link that can be opened on any device (e.g., smartphone, tablet, desktop computer). For our study, two types of mobile-friendly PROMs were collected from all patients over the course of the study: post-procedure recovery PROMs and disease-specific PROMs. Post-procedure recovery PROMs were emailed to patients twice weekly for a total of 5 weeks post-procedure. Bariatric surgery patients were asked two questions: 1) if they had pain (yes/no) and, if yes, to rate their pain on a scale of 1-10 and 2) if they had an appetite (yes/no) and, if yes, to rate their appetite on a scale of 1-10. Atrial fibrillation ablation patients were asked about pain, as well as whether they had palpitations and, if yes, to rate both symptoms palpitations on a scale of 1-10.

Disease-specific PROMs were emailed to patients at enrollment (pre-procedure) and 1, 4, and 8 weeks post-procedure and were tailored to patients depending on the procedure they received. For bariatric surgery patients, questions from the National Institute of Health’s (NIH) PROMIS® (Patient-Reported Outcomes Measurement Information System) relating to global health, pain, gastroesophageal reflux, nausea and vomiting, diarrhea, constipation, and sleep were adapted to a mobile-friendly format. For atrial fibrillation ablation patients, NIH PROMIS® questionnaires related to global health, dyspnea, and fatigue were adapted. Additionally, for these patients, we adapted the Cardiff Cardiac Ablation PROMS 1 and 2 to a mobile-friendly format.^10^ Beginning on May 21, 2018, approximately halfway through study completion, Hugo initiated a single weekly email reminder to patients who had not completed their PROMs within 24 hours.

For each PROM, the date and time that it was emailed to patients, the time it was initiated, and the time at which the final response was received were all available. Additionally, the content of response to each PROM question was obtained.

### Data Aggregation and Validation of EHR data Collected by Hugo

For each data source (EHR, pharmacy, digital device data, PROM), we determined the information made available via the Hugo platform. For EHR data, we validated the following components over 8 weeks follow-up: encounter date, encounter type, and encounter primary diagnosis. Specifically, we determined if encounters aggregated by Hugo matched encounters listed in each patient’s EHR (which were researcher-provisioned EHR views), and if there were any missing or discrepant encounters or diagnoses. We also examined reasons for discrepancies. Because of a complete change to the Mayo Clinic EHR partway through our study, our validations for Mayo Clinic patients were only performed in the new (Epic-based) EHR.

### Statistical Analysis

We conducted descriptive statistics with means and standard deviations or medians with interquartile range, as appropriate using Excel 2016 (Microsoft Corp.; Redmond, WA). For all patients, we characterized age and sex based on EHR data. We calculated the in-person total enrollment time (from the time the study coordinator met the patient, including informed consent, set up of Hugo and other accounts, and completion of the first disease-specific PROM). While all patients received specialty care for bariatric surgery or atrial fibrillation ablation at YNHH or Mayo Clinic, we also identified those patients who also receive primary care at each health system.

For data collected via personal digital devices, we examined the frequency with which patients used and synced their personal digital device data as well as the data elements obtained from each device and also examined these data by age and sex. We considered a patient to have used a device and contributed data if there was a recording for steps or sleep for Fitbit, weight for the Withings Body Scale, and an ECG reading for the Kardia Mobile at least once every 7 days during 8-weeks post-procedure. For personal digital devices, we also examined changes over time in activity (Fitbit), weight (Withings), and heart rate (Kardia Mobile). For patients with available pre- and post-procedure activity data, we compared each patient’s individual average pre- and post-procedure step count. For patients with available pre- and 8-week post-procedure weight data, we compared each patient’s individual average pre-procedure weight up to 7 days pre-procedure with the 8-week weight. For PROMs, we examined the completion rates, time to completion, as well as the proportion of survey items completed and completion rates by sex. For both personal digital device and post-procedure recovery PROM data, we examined trajectories in the content of data and responses over time.

Data from each personal digital device were plotted from 7 days pre-procedure (when available) and up to 8 weeks (56 days) post-procedure by calculating the mean steps, weight, and heart rate per day and stratified by the patient cohort using these devices. A polynomial best fit line was fitted to the data starting from day 0. Similarly, we plotted data from post-procedure recovery PROMs, by averaging the reported pain and appetite (for bariatric surgery patients) and pain and palpitations (for atrial fibrillation ablation patients) for each of the 10 surveys. For post-procedure recovery PROMs, the response score was counted as a “zero” when patients answered “no” to any of the questions.

## RESULTS

### Patient Demographics and Enrollment Data

Study coordinators screened a total of 77 patients between January 26, 2018 to October 19, 2018; 11 declined enrollment, 6 could not be enrolled as they did not meet the eligibility criteria, and 60 patients (30 patients at each site and 15 patients for each procedure) consented and enrolled, of whom 59 underwent the planned procedure (one patient scheduled for bariatric surgery did not receive the procedure). The mean patient age was 55.2 years (standard deviation 13.5). Overall, 58.3% (n=35) of the 60 patients enrolled were female, including 76.7% (n=23) of the 30 bariatric surgery and 40.0% (n=12) of the 30 atrial fibrillation ablation patients. The median in-person enrollment time was 70 minutes (interquartile range [IQR]: 58 to 100); 69 minutes (IQR, 46-76) for bariatric surgery patients and 73 minutes (IQR, 50-93) for atrial fibrillation ablation patients, with 7 patients completing some enrollment steps remotely. The final patient completed follow-up as of December 25, 2018.

### EHR Data: Aggregation

EHR data were successfully aggregated from both the Yale-New Haven Hospital (YNHH) and Mayo Clinic EHRs for all patients who underwent their procedure. Among the 59 patients, 32 (54%) also received primary care services at YNHH or Mayo Clinic, in addition to the specialty care that led them to undergo either bariatric surgery or atrial fibrillation ablation at the hospital. Of the 51 patients reached to complete close-out surveys, 18 (35%) reported receiving care at an institution other than YNHH or Mayo Clinic during the follow-up period; 6 (33%) of these 18 patients connected to outside EHR portals. Of the 27 patients who did not receive primary care at YNHH or Mayo Clinic, 10 (37%) used Hugo to link their EHR portals from other health systems. In total, data from 13 EHRs were aggregated by Hugo for the 59 patients. During the study, the Hugo platform added new connections to three additional portals after patients identified receiving care at those health systems.

### EHR Data: Validation

Among the 59 patients, there were 271 encounters identified in either the EHR data or Hugo, 221 (82%) were observed in both sources, 36 (13%) were observed only in the EHR data, and 14 (5%) were observed only in Hugo. For 5 patients (4 at YNHH and 1 at Mayo Clinic), EHR data did not sync for the entire follow-up period, accounting for all 36 encounters observed in the EHR data but not in Hugo; the most common reason for sync failure was that the portal account was locked after multiple incorrect password entries (by the patient, not by Hugo). Among the 14 encounters observed only in Hugo, 6 (43%) were among YNHH patients and 8 (57%) were among Mayo Clinic patients. These 14 encounters occurred at facilities or clinics within or affiliated with the health center, such that data representing these visits are included in the Continuity of Care document (CCD) and patient portal but are generally not populated within the health system EHRs because of patient privacy concerns (e.g, mental health encounters).

Of the 221 encounters observed in both the EHR data and in Hugo, the encounter date was identical for 215 (97%), the encounter type for 203 (92%), and the primary diagnosis for 204 (92%) (Table). One encounter date was missing (a mental health encounter) and one was a single day off. For encounter types with discrepancies, these were usually because the description was more specific in the EHR (e.g., “lab” or “imaging test”) than the description aggregated by Hugo (e.g., “outpatient” or “inpatient”). Two encounters were listed as “inpatient” within the EHR but “outpatient” in Hugo and one was listed as “outpatient” in the EHR but “inpatient” in Hugo. For diagnoses with discrepancies, nearly all occurred after a major YNHH Epic upgrade and were missing in Hugo; this upgrade required additional mapping from the Hugo platform to the YNHH patient portal. A single encounter diagnosis was listed as “paroxysmal atrial fibrillation” in EHR, “cardiac arrhythmia, unspecified cardiac arrhythmia type” in Hugo.

**Table:** Data in Hugo and Concordance with Yale New Haven Health and Mayo Clinic Epic Electronic Health Record Data for Encounters Found in Both Data Sources

| Data from Yale New Haven-Health and Mayo Clinic Epic-based EHRs | Data Obtained from Hugo That are Concordant with EHRs<br>N (% concordant) |  |  |
| --- | --- | --- | --- |
|  | Encounter Date | Encounter Type | Primary Diagnosis |
| **Atrial Fibrillation Ablation**<br>**n=115 encounters** | 110 (96%) | 100 (87%) | 101 (88%) |
| **Bariatric Surgery**<br>**n=106 encounters** | 105 (99%) | 103 (97%) | 103 (97%) |
| **Overall**<br>**n=221 encounters** | 215 (97%) | 203 (92%) | 204 (92%) |

### Pharmacy Data: Aggregation

Among the 59 patients, 24 (41%) stated that they obtained at least some of their medications at either CVS or Walgreens pharmacies and connected their accounts to Hugo. Patients who did not use CVS or Walgreens either used Walmart, a large pharmacy that at the time of the study lacked an established API/patient portal, or smaller local pharmacies, mail order pharmacies, or grocery store pharmacies. Among the 15 patients with available CVS or Walgreens medication data, the mean number of prescriptions orders over the 8-week follow-up period was 4.5 (SD 4.3).

### Personal Digital Devices: Aggregation

For all 59 patients, weekly Fitbits syncs decreased from 47 patients who synced (80%) at week 1 to 34 patients (58%) at week 8; 64% overall for the study period (Figure 1). A total of 32 patients (54%) synced their Fitbit across all 8 weeks. Of the 118 total Fitbit syncs at 1-, 4-, and 8-weeks post-procedure, step data were available for 114 (97%) and sleep data for 99 (84%). Among the 29 patients who underwent bariatric surgery, weekly Withings Body scale syncs decreased from 20 patients (69%) at week 1 to 14 patients (48%) at week 8; 56% overall for the study period. Eleven (38%) of the 29 bariatric surgery patients recorded a weight for all 8 weeks post-procedure. All 131 Withings syncs included data on patient weight. Among the 30 patients who underwent atrial fibrillation ablation, 20 (67%) synced their Kardia Mobile at both weeks 1 and 8; 70% overall for the study period, as the specific patients syncing changed over time. Fifteen (50%) of the ablation patients recorded a Kardia Mobile ECG for all 8 weeks. All 1,340 Kardia Mobile syncs included data on reading duration, heart rate, and rhythm interpretation.

Examining weekly Fitbit syncs by age, 75%, 66%, and 52% of patients in the 18-44, 45-64, and 65+ age groups, respectively, synced their device, whereas for the second device (weight scale or Kardia Mobile), rates were 73%, 59%, and 61%, respectively. Examining weekly Fitbit syncs by sex, women completed 67% and men 60%. For the second device, these values were 58% and 69%, respectively.

### Personal Digital Devices: Patterns of Recovery

Personal digital device data from patients’ Fitbits demonstrated that, on average, patients in both bariatric surgery and atrial fibrillation ablation groups increased their daily step counts in the first several days post-procedure and then plateaued (Figures 2 and 3). Among the 22 bariatric surgery patients and 21 atrial fibrillation ablation patients with available pre-procedure data, 13 (59%) and 14 (66%), respectively had higher post-procedure average step counts when compared to each individual’s pre-procedure step count. Personal digital device data from Withings scales demonstrated that among patients who recorded both a pre-procedure and week 8 weight, the median weight loss was 16.1 kilograms (IQR, 15-19) (Figure 4). Data from Kardia Mobile showed that of 1,340 readings by 30 patients, 834 (62%) detected normal sinus rhythm, 228 (17%) detected possible atrial fibrillation, 120 (9%) were an undetermined rhythm, and 158 (12%) had no definitive rhythm determination due to factors such as background noise or premature device disconnection. Average heart rate data indicate a small increase during the first 30 days and then a decrease (Figure 5).

### PROMs: Aggregation

Among the 59 patients, 440 of 539 (82%) post-procedure recovery PROMs were completed; 203 of 260 (78%) among the 29 bariatric surgery patients and 237 of 279 (85%) among the 30 atrial fibrillation ablation patients (Figure 6). Rates of completion were 80%, 81%, and 84%, among patients in the 18-44, 45-64, and 65+ age groups, respectively, whereas women completed 83% and men 79%. On average, bariatric surgery patients completed 7.1 of 10 (71%) post-procedure recovery PROMs they received and atrial fibrillation ablation patients completed 7.7 of 10 (77%) post-procedure PROMs; 57 (97%) patients completed at least one post-procedure recovery PROM. The median time spent responding to post-procedure recovery PROMs per week was 20 seconds (IQR, 13-31), 14 seconds (IQR, 9-23) for bariatric surgery patients and 22 seconds (IQR, 17-41) for atrial fibrillation ablation patients.

For disease-specific PROMs, 123 of 148 (83%) were completed; 64 of 75 (85%) among the bariatric surgery patients and 59 of 73 (81%) among the atrial fibrillation ablation patients (Figure 6). Rates of completion were 86%, 85%, and 77%, among patients in the 18-44, 45-64, and 65+ age groups, respectively, whereas women completed 85% and men 81%. On average, bariatric surgery patients completed 3.2 of 4 (80%) disease-specific PROMs they received, atrial fibrillation ablation patients 2.7 of 4 (68%); 52 (88%) completed at least one disease-specific PROM. The median time spent responding to disease-specific PROMs was 8 minutes (IQR, 5-12), 5 minutes (4-7) for bariatric surgery patients and 11 minutes (8-16) for atrial fibrillation ablation patients. Of the 123 disease-specific PROMs completed, 46 (37%) were 100% complete and 118 (96%) included responses to at least 90% of the survey questions.

### PROMs: Patterns of Recovery

PROM data reported by patients who underwent bariatric surgery demonstrated that appetite levels were initially low for the first week, and then increased and remained moderate and generally steady during the 5-week post-procedure period (Figure 7) during which post-procedure recovery was assessed, whereas pain decreased over the first 1-2 weeks and then generally plateaued except for a slight increase at weeks 3-4. Similarly, PROM data reported by patients who underwent atrial fibrillation ablation demonstrated that palpitations decreased slightly over the first two weeks but then increased at week 3 (Figure 8) and then decreased again, whereas pain levels remained low during the post-procedure period.

## DISCUSSION

Our study demonstrates the feasibility of using the Hugo sync-for-science platform to obtain and aggregate patient data from multiple real-world data sources, including EHRs, pharmacies, PROMs, and personal digital devices, over an 8-week follow-up period in a near real-time, streaming fashion for patients who underwent atrial fibrillation ablation or bariatric surgery. These results suggest that there is great potential in using novel digital health technologies to enable aggregation of patient data across multiple sources for research and regulatory purposes. While our study was focused on procedures that use medical devices, the principles could be used for evaluating the safety and effectiveness of any medical product, as well as for better understanding patient recovery, function and symptoms after undergoing any medical procedure or surgery, behavioral intervention, or even a health care delivery redesign.

The FDA is increasingly using real-world evidence for medical product evaluations, including both medical devices and pharmaceuticals. The National Evaluation System for Health Technology (NEST) has been created to support this goal, with the mission of accelerating development and translation of new and safe health technologies by leveraging real-world evidence and innovative research.^11,12^ Through NEST, a comprehensive and accurate framework can provide information about risks and benefits of devices.^12^ Similarly, for pharmaceuticals, FDA recently released a Framework for its Real-World Evidence Program, pursuant to the 21^st^ Century Cures Act, for the potential use of real-world evidence to support approval of a new indication for a drug on the market or to help support or satisfy post-approval requirements. These goals rely on obtaining and aggregating data from increasingly available real-world data sources, such as personal digital devices or digital PROMs. More importantly, it depends on removing the sequestration of data sources (including sequestration within a single source, such as EHRs lacking interoperability) and integrating them with other sources to provide a more comprehensive understanding of medical product performance and patient outcomes. Aggregation of multiple data sources allows data validation and overcomes the unreliability of a single data source.^13,14^ Our study shows the potential to stream near real-time data during post-procedure follow-up from multiple digital sources and integrate them into a single dataset for research purposes, akin to a pragmatic clinical cohort study that required only upfront time and effort to enroll patients in the Hugo platform, but which otherwise relied on passive data collection without requiring patient visits with study personnel.

Continuous digital health technology advances are likely to help support the realization of this potential. For example, after initiation of our study, the Hugo platform made updates expected to improve response rates to PROMs and personal digital device data syncs, including text messaging reminders (this study used email reminders only) and text messaging PROMs with the goal of improving future workflows. For some patients, response rates may be higher through use of text messaging or app notifications compared to email. Data from patient APIs are also expected over time, providing additional, detailed information. Further, the gradual, increasing penetration and individuals’ facility of digital technology, use of PROMs, personal digital devices, and additional sources are likely to increase patient engagement. In particular, personal digital devices, such as consumer wearables and sensors, are generating data previously unused in routine clinical care. Connecting these data to cloud-based sources will improve data aggregation and response rates, so that users will not be required to sync their devices to a connected interface to upload information. All of these advances will strengthen the potential for evaluations of patients being treated with devices, pharmaceuticals, and procedures through the use of multiple real-world data sources by improving understanding of patient recovery, function, and symptoms.

Our study is notable for its patient-centered nature: patients received access to their data and then were asked to contribute it to advance science and research. People continue to obtain greater access to their health data, such as through Medicare’s Blue Button 2.0, which enables over 50 million beneficiaries to obtain access to and share their claims data.^15^ Health systems and pharmacies are also making more health data available to patients, including clinician notes;^16^ as data are increasingly made directly available to patients, they will be able to share their own data for research purposes or use the personal health record available on their mobile device to provide additional information to their treating physicians for clinical purposes. Through aggregating multiple data sources and enabling patients to share their data, evaluations of medical products, patient status, and patient recovery can now be performed; thus far, such research has been difficult in the current limited data environment. This may be increasingly important as complex care is more frequently provided at tertiary medical centers, which may require patients to travel to obtain access to procedures. Preoperative lack of information along with post-operative loss to follow-up may bias outcomes research. One current limitation of using these data for medical device evaluations is that the unique device identifier, which allows identification of any medical device, has not been integrated into EHRs; accomplishing this integration and then providing the information to patients will strengthen the ability to study medical device safety and effectiveness.^17^

Our study should be considered in the context of important limitations. First, as this was a feasibility study, we enrolled a small number of patients (60) for a relatively short follow-up duration (8 weeks). However, there is no reason to believe that our findings would not scale to a larger number of diverse patients and for a longer follow-up duration. Further, many device-related events occur peri-procedurally and in the immediate post-procedure follow-up (particularly for atrial fibrillation ablation)^18^ and, thus we would expect to capture them during this follow-up period. As no patient reported intentionally stopping data sharing to our knowledge, we expect longer follow-up durations would have similar success in obtaining data. Moreover, this study demonstrates the importance of ‘data stream checks’ to ensure that data aggregation has not been interrupted. Second, we provided stipends to cover time and effort involved with study enrollment and participation and free personal digital devices, both of which may have enhanced engagement over the follow-up period. However, many patients reported being motivated to participate because they wanted to share their data with their clinicians; the opportunity to have their data integrated with clinical care is likely to be a major motivation, particularly given the association with improved outcomes.^19,20^ Third, we did not examine the utility of all the data obtained; because of variability in data availability, additional processing will likely be necessary to use various data elements for medical product evaluations.

In conclusion, our study demonstrated the feasibility of using a novel patient-centered health data sharing platform, Hugo, to aggregate multiple real-world data sources in near real-time, thereby enabling evaluation of medical products, in this case medical devices, for both research and regulatory purposes and facilitating better understanding of patient recovery, function and symptoms.

## Data Availability

Data Availability: The dataset generated and analyzed for this study will not be made publicly available due to patient privacy and lack of informed consent to allow sharing of patient data outside of the research team.

## Data Availability

The dataset generated and analyzed for this study will not be made publicly available due to patient privacy and lack of informed consent to allow sharing of patient data outside of the research team.

## Acknowledgments

This work was supported in part by a Center of Excellence in Regulatory Science and Innovation (CERSI) grant to Yale University and Mayo Clinic from the US Food & Drug Administration (U01FD005938) and by Johnson & Johnson. Its contents are solely the responsibility of the authors and do not necessarily represent the official views nor the endorsements of the Department of Health and Human Services, FDA, or Johnson & Johnson. We gratefully acknowledge the National Evaluation System for Health Technology (NEST) designation of this work as a Demonstration Project and the Hugo team for their assistance throughout this project. We acknowledge AliveCor for their generous donation of the Kardia Mobile devices used in this study. Most importantly, we gratefully acknowledge the contributions of patients to this study.

## Author Contributions

S.S.D., J.S.R., and N.D.S. contributed to the conceptualization and design of the study. J.G.A, B.C., K.C., W.C., L.C., H.J.D., J.D., S.J., T.K., C.L., P.A.N., K.R., A.S., A.Y., and N.D.S. contributed to the development of the study protocol. J.G.A., T.K., P.A.N., K.R., and cardiac electrophysiologists and bariatric surgeons at Mayo Clinic identified patients for enrollment. L.C. and H.J.D. oversaw patient recruitment and performed the data validation and analysis. All authors contributed to data interpretation. L.C. performed descriptive statistical analyses and created the data visualization, figures, and table. S.S.D., J.S.R., L.C., H.J.D., and N.D.S. wrote the manuscript. All authors critically reviewed the manuscript.

## Additional Information

### Potential Competing Interests (past 36 months)

Dr. Dhruva currently receives research support through the National Institute of Health (K12HL138046). Dr. Ross formerly received research support through Yale University from Medtronic, Inc. and the Food and Drug Administration (FDA) to develop methods for postmarket surveillance of medical devices (U01FD004585), from the Centers of Medicare and Medicaid Services (CMS) to develop and maintain performance measures that are used for public reporting (HHSM-500-2013-13018I), and from the Blue Cross Blue Shield Association to better understand medical technology evaluation; Dr. Ross currently receives research support through Yale University from Johnson and Johnson to develop methods of clinical trial data sharing, from the Medical Device Innovation Consortium as part of the National Evaluation System for Health Technology (NEST), from the Agency for Healthcare Research and Quality (R01HS022882), from the National Heart, Lung and Blood Institute of the National Institutes of Health (NIH) (R01HS025164), and from the Laura and John Arnold Foundation to establish the Good Pharma Scorecard at Bioethics International and to establish the Collaboration for Research Integrity and Transparency (CRIT) at Yale. In the past 36- months, Dr. Shah has received research from the Centers of Medicare and Medicaid Innovation under the Transforming Clinical Practice Initiative (TCPI), from the Agency for Healthcare Research and Quality (R01HS025164; R01HS025402; 1U19HS024075; R03HS025517), from the National Heart, Lung and Blood Institute of the National Institutes of Health (NIH) (R56HL130496; R01HL131535), National Science Foundation, and from the Patient-Centered Outcomes Research Institute (PCORI) to develop a Clinical Data Research Network (LHSNet). Drs. Coplan and Yoo, Mr. Johnston, and Ms. Childers are employees and stockholders of Johnson & Johnson; Dr. Chow is an employee of Janssen Scientific Affairs, LLC, and a stockholder of Johnson & Johnson.

